# Absolute quantification and degradation evaluation of SARS-CoV-2 RNA by droplet digital PCR

**DOI:** 10.1101/2020.06.24.20139584

**Authors:** Curtis J. Mello, Nolan Kamitaki, Heather de Rivera, Steven A. McCarroll

**Affiliations:** Department of Genetics, Harvard Medical School; Broad Institute of MIT and Harvard

## Abstract

Quantifying SARS-CoV-2 infectivity, formulating well-calibrated public-health policy, and managing the safety of workplaces would all be facilitated by precise measurement of the extent to which SARS-CoV-2 RNA is present in an intact form in biological specimens and human environments. We describe assays that use digital PCR in nanoliter droplets (droplet digital PCR) to measure these properties. Such assays could be broadly deployed to inform COVID-19 epidemiology, measure symptomatic and asymptomatic infectivity, and help manage the safety of environments in which people live, move, and work.

## Background

The ongoing COVID-19 pandemic is caused by SARS-CoV-2, a positive-sense single-stranded RNA virus. The SARS-CoV-2 genome consists of a single linear RNA segment ∼30,000 bases in length, that generates multiple subgenomic RNA transcripts^1,2^. Many important medical, organizational, personal and public-health decisions hinge on key variables whose values are still unknown, including the extent to which SARS-CoV-2 is present in respiratory droplets at pre-symptomatic, symptomatic, and convalescent stages of illness and in infected individuals who never manifest symptoms^3,4^. For example, planning for the reopening of schools would ideally be informed by information about how viral shedding varies as a function of age, detectable symptoms, and time since exposure, all of which remain substantively unknown^5-8^. Measuring these parameters will be critical in predicting infection trajectories and determining the adequacy of workplace safety measures and shift schedules. Similarly, in managing the resumption of workplace activity, it will be critical to assess the extent to which SARS-CoV-2 is present in human environments, including public transportation and high-contact surfaces. Such decisions will need to be revised and adapted in a dynamic way that incorporates emerging data that are precise, quantitative, and informative.

Diagnostic assays for the presence of SARS-CoV-2 currently use real-time reverse transcriptase PCR (RT-qPCR) to yield a binary (positive or negative) result based on an amplification cycle threshold (Ct) value^9-12^. However, Ct values are shaped by laboratory-specific and instrument-specific variables and require calibration against an uncertain standard sample^13,14^. Hence, there is a need for assays that measure absolute quantities of viral material and are comparable across laboratories.

It is also important to distinguish intact SARS-CoV-2 genomes from degraded viral RNA, as the latter contribute to positive test results but are an uncertain proxy for infectiousness. The gold standard measurement for infectiousness of a sample, a cell culture plaque assay to determine viral titer, must be performed in a biosafe facility with specialized equipment for culturing infectious virus, requires established cell lines that can host SARS-CoV-2, and takes days^15^. Together, these constraints limit throughput and create obstacles to performing these measurements on samples collected at many locations. Current PCR-based assays can detect the presence of very short SARS-CoV-2 RNA sequences but do not distinguish whether these sequences are derived from longer molecules present in the sample at the time of collection. Degraded RNA may linger in human environments despite successful decontamination efforts and might also remain detectable in once-infected individuals long after recovery and conversion to a non-infectious status^5,6,16-18^. The observed tendency of recovered individuals to fluctuate between positive and negative results has created uncertainty about the durability of adaptive immunity, limited the contribution of convalescent plasma for passive immunity therapy, and increased strain on hospital facilities by limiting discharge. It is important to understand whether such fluctuations reflect truly relapsed infection and infectiousness, or temporal variation in the levels of degraded viral nucleic acids that can be detected by assays.

To address these issues, we explored using droplet digital PCR (ddPCR)^19,20^ to more precisely quantify SARS-CoV-2 RNA in biological samples and evaluate the extent to which positive results reflect larger, intact viral nucleic acids. In ddPCR, a liquid biological sample is distributed, along with enzymes, nucleotides, and oligonucleotides for reverse transcription and PCR, into tens of thousands of nanoliter-sized aqueous droplets in an oil-aqueous emulsion. Reverse transcription and PCR are performed within all droplets simultaneously; dual-labeled (fluorophore/quencher) oligonucleotide probes fluoresce upon template amplification and consequent probe cleavage; and fluorescent (i.e., template-positive) droplets are digitally counted as they flow through microfluidic devices, allowing digital counting of the number of template molecules per unit volume in the input sample. The use of digital counting improves the quantitation of template nucleic acid sequences relative to real-time PCR, yielding measurements with a clear biological interpretation that can be readily compared across biological samples and laboratories.

Droplets also make it possible to determine whether distinct sequences are present on the same larger nucleic acid (e.g., an intact viral genome). The principle of such analysis is that when distinct nucleic acid sequences are physically linked, they will be in the same droplets, whereas unlinked sequences will co-encapsulate in droplets (by chance) at a far lower rate. We previously described the application of such an approach to determine the chromosomal phase of human DNA sequence variants^21^. Here we apply this idea to measure the integrity of SARS-CoV-2 RNA.

## Materials and methods

### Probes and primers

Initially, we used the published primer and probe sequences designed for CDC’s 2019-nCoV Real-Time RT-PCR Diagnostic Panel. This panel includes the primer/probe sets N1 and N2, which have been approved for testing clinical samples for the presence of SARS-CoV-2 RNA by real-time RT-qPCR. The N1 and N2 assays detect two separate locations about 1 kb apart that are specific to the SARS-CoV-2 nucleocapsid (N) gene sequence. Both the probe assays distributed through the Emergency Use Authorization (EUA) for clinical diagnostics and commercially available Research Use Only (RUO) kits are labeled with a 6-carboxyfluorescein (FAM) molecule and a quencher; consequently, the assays must currently be run as separate reactions. To couple the two probes into a single ddPCR reaction, we ordered the N1 and N2 assays from Integrated DNA Technologies (IDT), with each probe labeled at the 5’ end with either fluorescein (FAM) or hexachlorofluorescein (HEX) dyes. The sequences of the N probe assays are available at https://www.cdc.gov/coronavirus/2019-ncov/lab/rt-pcr-panel-primer-probes.html.

We then designed a ddPCR probe assay targeting the ORF1a region of the SARS-CoV-2 genome using the program Primer3Plus and the complete SARS-Cov-2 genome (GenBank Sequence Accession MN908947) The sequences chosen for the ORF1a assay are as follows: fwd. primer, CGTAGTGGTGAGACACTTGG; reverse primer, TACGAAGAAGAACCTTGCGG; Probe (FAM): TGGGCGAAATACCAGTGGCT.

We assembled a 20× Probe/Primer mix for each FAM or HEX assay by reconstituting each lyophilized component from IDT to 100 µM, and then combining: 5.5 µL probe; 19.8 µL forward primer; 19.8 µL reverse primer; 64.9 µL dH_2_O (volumes can be scaled up). This 20× master mix can be frozen and used in subsequent experiments.

### Positive control

Commercially available SARS-CoV-2 RNA standard from Exact Diagnostics (cat #COV019), which contains RNA transcripts for the SARS-CoV-2 ORF1a and N genes (among others). We note that the use of this preparation as a positive control does not address potential challenges in obtaining an adequate specimen from a patient and extracting viral RNA intact from this sample.

### Negative control

RNA extracted from HEK 293 (human embryonic kidney) cells.

### Droplet digital PCR (ddPCR)

Each ddPCR assay reaction (22 µL total volume) was set up on ice using reagents from the One-Step RT-ddPCR Advanced Kit for Probes (Bio-Rad #1864020): per reaction, 5.50 µL One-Step Supermix; 2.20 µL Reverse Transcriptase; 1.10 µL of 300 mM DTT; 1.21 µL of 20X FAM Probe/Primer mix; 1.21 µL of 20× HEX Probe/Primer mix; 9.78 µL dH2O; and 1.00 µL sample (10–50 ng/µL). (Note that the reagents other than the test sample are ideally made into a larger master mix and then aliquoted into individual reactions.)

Twenty microliters of the sample mix was loaded into a Bio-Rad QX200 droplet generator with 70 µL Droplet Generation Oil for Probes, as specified by the manufacturer. Forty microliters of droplets were recovered per reaction. Thermal cycler conditions were as follows: 50°C for 60 min; 95°C for 10 min; 50 cycles of 95°C for 30 sec and 60°C for 60 sec; 98°C for 10 min; 12°C hold.

### Quantitative calculation of linkage

The absolute number and fraction of molecules on which the detected sequences are physically linked was calculated according to a formula we derived and described in earlier work (see Note S1 of ref ^21^). This quantity is calculated from the numbers of double-positive, FAM-positive, HEX-positive, and double-negative droplets; it does not exactly match the number (or proportion) of double-positive droplets because some double-positive droplets are assumed to be due to chance co-encapsulation.

## Results

### Physical linkage of SARS-CoV-2 RNA sequences evaluated in droplets

To investigate whether ddPCR could be used to determine whether two SARS-CoV-2 RNA sequences are physically linked, we performed two experiments. In the first, we simultaneously detected two sequences derived from the SARS-CoV-2 N gene. A mixture of SARS-CoV-2 template RNAs, including RNAs containing the N and ORF1a genes, were encapsulated into droplets with two primer-probe sets corresponding to different sequences from the SARS-CoV-2 N gene (N1 and N2, the same sequences used in the CDC-approved assay). These assays detect sequences that are separated by 806 bases on the SARS-CoV-2 genome and on transcripts of the N gene (**Fig. 1**).

**Figure 1.**
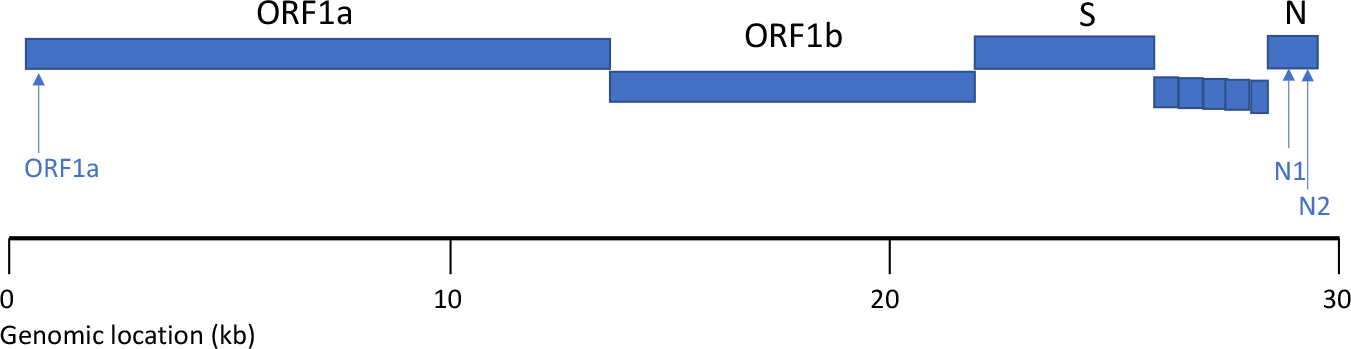
Sites on the SARS-CoV-2 genome and RNA transcripts detected by the primer/probe sets in this work.

The results yielded definitive evidence of linkage: although only 822 of the 12,220 droplets (6.7%) were positive for either N1 or N2, 75% of the droplets that were positive for N2 (HEX) were also positive for N1 (FAM) (**Fig. 2a, Table 1**); we estimate (using a formula we previously described^21^, which accounts for chance co-encapsulation) that 71% of the detected RNA sequences were physically linked.

**Figure 2.**
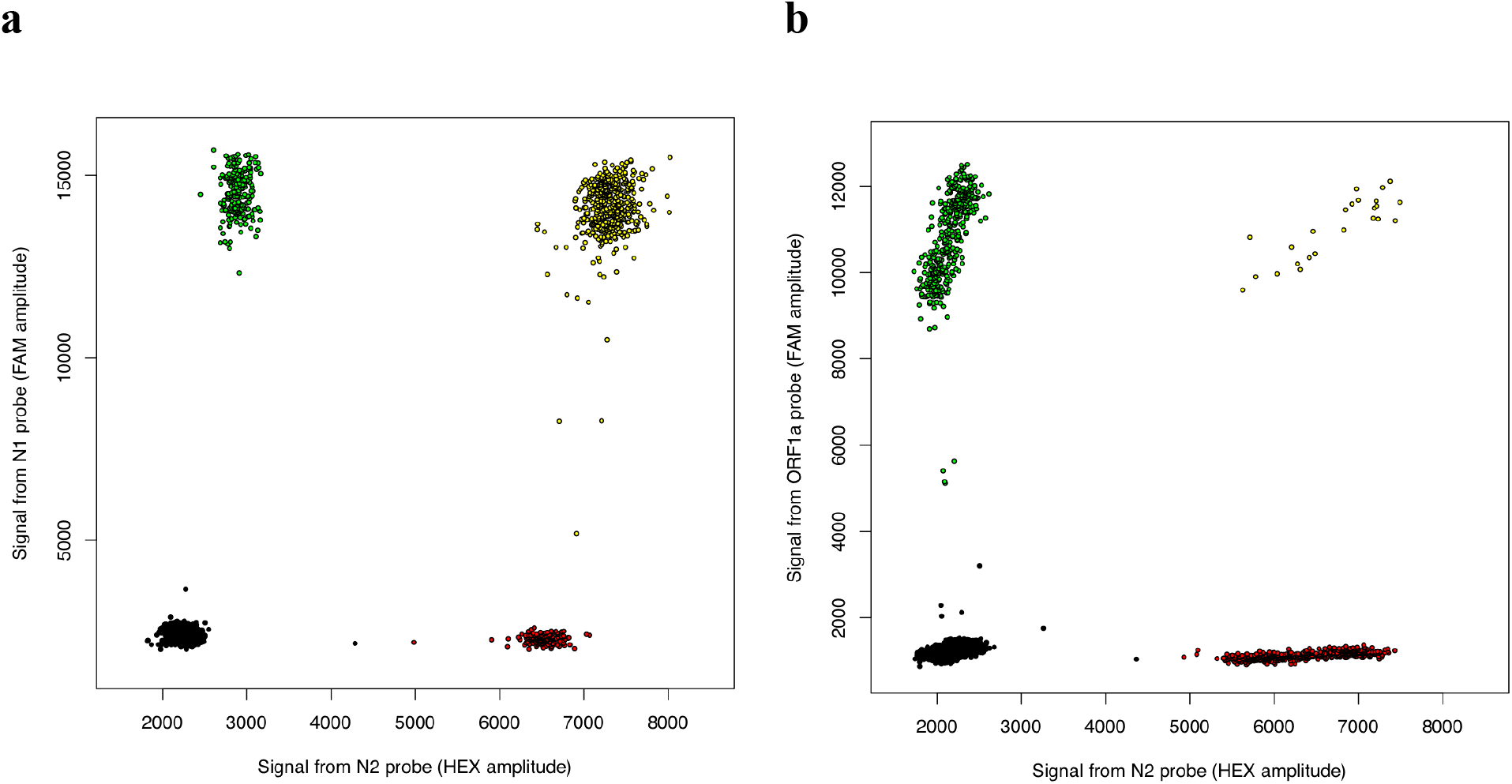
Droplets distinguish between physically linked and unlinked SARS-CoV-2 RNA sequences. **(a)** Two sequences derived from the N gene were most frequently detected in the same droplets. **(b)** Sequences derived from the N and ORF1a genes were generally detected in distinct droplets, co-localizing only to the extent expected by chance. Droplets determined to be FAM+HEX+ (i.e. to have contained both templates) are shown in yellow; FAM+HEX-droplets in green; FAM-HEX+ droplets in red; and FAM-HEX-droplets in black.

**Table 1.**
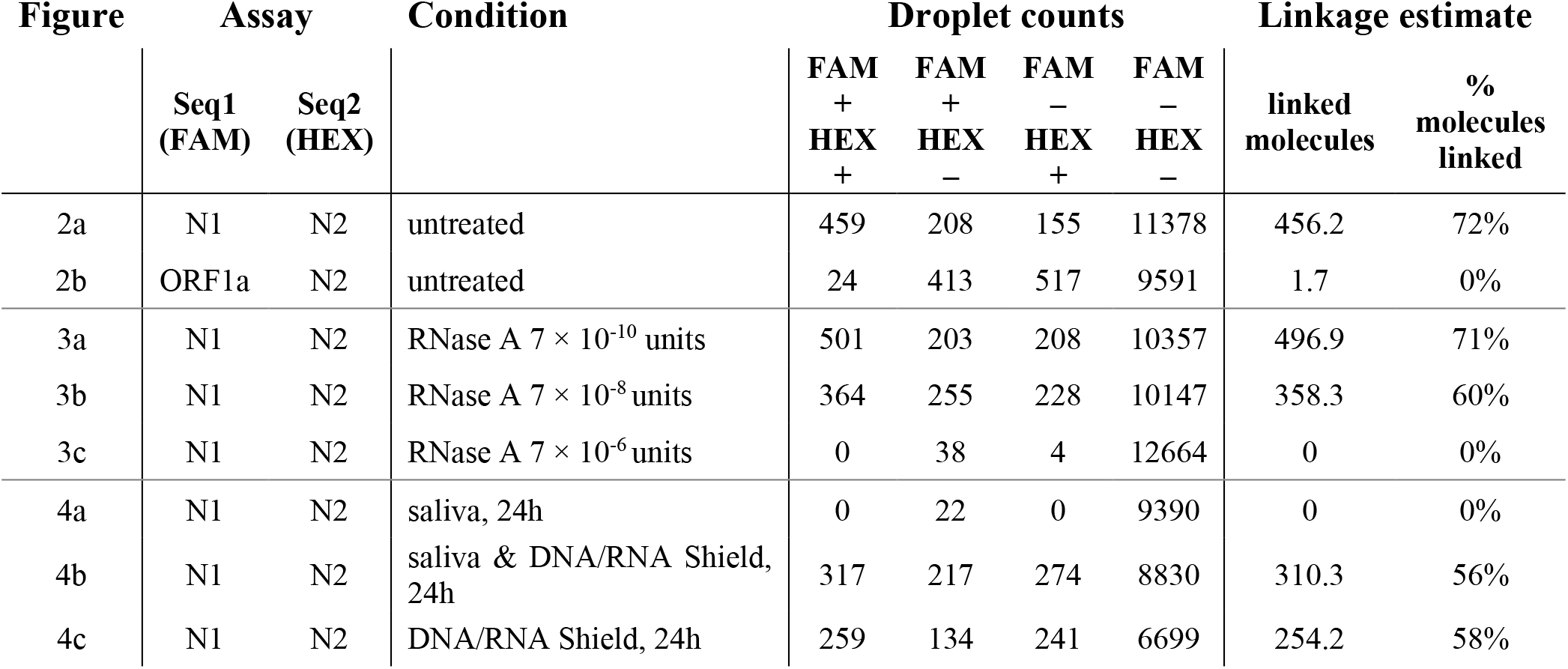
Digital counts of droplets positive and negative for the two template sequences in each assay, and estimates (calculated from these droplet-count data) of the number and percent of RNA molecules on which these sequences are physically linked. Linkage estimates were calculated from the primary droplet data using a formula we described in earlier work (see Note S1 of ref^21^). Note that counts for two sequences present at equimolar concentrations will differ due to molecular sampling and can also differ due to differences in target accessibility or fragmentation; the estimate of the percent of molecules on which these sequences are linked therefore utilizes an average measured concentration of the two assay sequences.

Next, we encapsulated a SARS-CoV-2 RNA mixture with primer–probe pairs detecting sequences on two distinct RNAs: the ORF1a transcript (detected with a FAM-labeled probe) and the N gene (detected with a HEX-labeled probe) (**Fig. 1**). Although both sequences were robustly detected, droplets were double-positive only to the extent expected based on chance co-encapsulation of physically unlinked molecules (**Fig. 2b**). Specifically, 4–5% of droplets were positive for N; 4– 5% were positive for ORF1a; and of those droplets that were positive for either template, 4–5% were positive for both templates, consistent with the absence of linkage (**Fig. 2b, Table 1**).

### Degradation of SARS-CoV-2 RNA sequences in proportion to RNase exposure

To evaluate the ability of ddPCR to quantify degradation of SARS-CoV-2 RNA sequences, we exposed the same SARS-CoV-2 RNA templates to RNase A (Qiagen #19101) at a series of concentrations. 15 µL of template RNA was combined with 1 µL of diluted RNase A and incubated at 37°C for 10 minutes, then 3 µL of RNase Inhibitor (Lucigen #30281) was added. At the lowest tested RNase A concentration (7 × 10^−10^ units), most N1 and N2 sequences were still detected in the same droplets, yielding an estimate of 71% linked molecules (**Fig. 3a, Table 1**), similar to the no-RNAse condition (72%, **Table 1**). When RNA templates were exposed to less-dilute RNase (7 × 10^−8^ units), linkage was reduced to 59% (**Fig. 3b, Table 1**). At a higher RNase concentration (7 × 10^−6^ units), detection of the individual N1 and N2 sequences was also greatly reduced, and no linked molecules were detected (**Fig. 3c, Table 1**).

**Figure 3.**
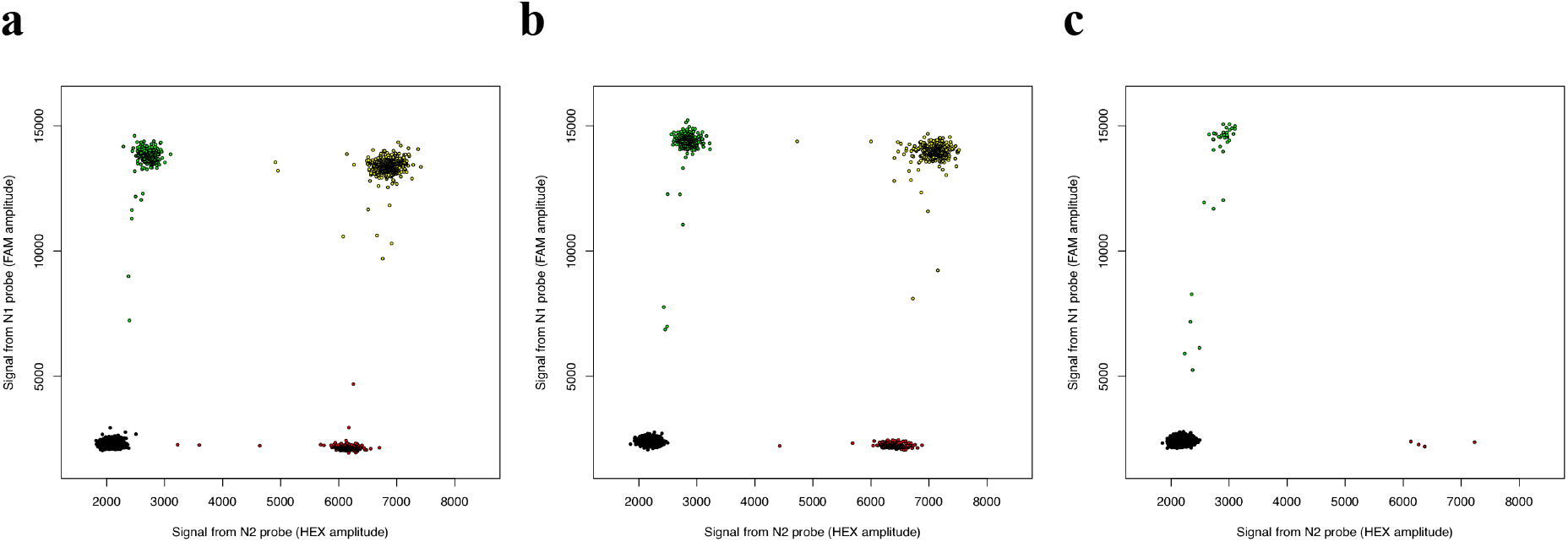
Concentration and physical linkage of SARS-CoV-2 RNA sequences was measured by ddPCR after exposure to RNase A at concentrations of (**a**) 7 × 10^−10^ units, (**b**) 7 × 10^−8^ units, (**c**) 7 × 10^−6^ units, yielding estimates of 53%, 39%, and 0% linkage respectively. Points colored as in Fig. 1.

### Preservation and detection of SARS-CoV-2 RNA sequences in saliva

Detection of SARS-CoV-2 in saliva may offer an enabling alternative to the current practice of detecting SARS-CoV-2 from nasopharyngeal swabs^22^. The current scarcity of swabs limits the availability of testing even for diagnostic purposes. Moreover, saliva can be obtained from patients in practices and contexts that do not put health care workers at risk, potentially even including home testing. Saliva tests for SARS-CoV-2 are also potentially more consistent and sensitive than swab tests, as the administration of swabbing may be inconsistent across patients and sessions, whereas concentrations of viral RNA in saliva can be normalized volumetrically. Critical for enabling widespread saliva testing in homes and workplaces is the ability to collect saliva into media that preserve viral RNA for analysis.

Saliva contains abundant nucleases that might reduce the detectability of SARS-CoV-2 templates. To test this possibility, we incubated SARS-CoV-2–derived RNAs with human saliva at room temperature for 24 hours. Under these conditions, N2 sequences were no longer detected, and N1 sequences were detected at a low level (< 1% of their initial concentration) (**Fig. 4a, Table 1**) that was also observed in a zero-template negative control sample and may thus represent a low level of contamination (data not shown). (We note that this observation does not reflect or predict the survivability of intact, infectious viral particles inside of saliva droplets.) When the same sample (saliva + SARS-CoV-2–derived RNA) was mixed with an RNA stabilization reagent (DNA/RNA Shield, Zymo Biosciences) and incubated at room temperature for 24 hours, our ddPCR assays robustly detected both sequences and detected strong linkage (est. 56% linked molecules) between these sequences (**Fig. 4b, Table 1**), similar to the level of linkage (58%) was detected in a control RNA sample that was not exposed to saliva (**Fig. 4c, Table 1**).

**Figure 4.**
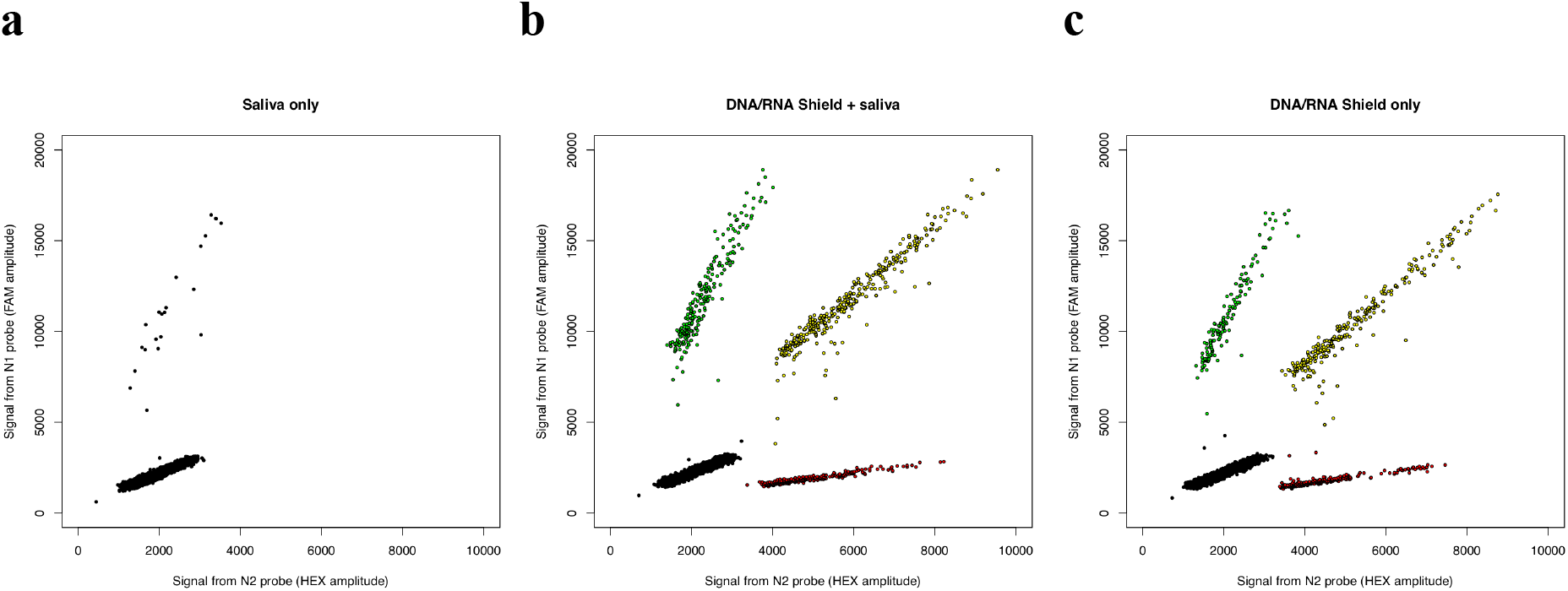
Analysis by ddPCR of concentration and linkage (co-partitioning) of two SARS-CoV-2-derived RNA sequences (**a**) when viral RNA was pre-incubated with human saliva; (**b**) when a virus-inactivating, nucleic acid– preserving medium was added at the start of incubation with saliva (room temperature for 24 hours); and (**c**) in the absence of saliva, with the preservation medium alone. (Note that heterogeneity in droplet size or quality can cause the droplet clusters to be diagonally oriented.)

## Conclusions

The assays we have described here could be readily established in diverse laboratories, at modest reagent cost (about $10 per sample) and in a 96-well format, using widely available instruments to perform ddPCR. The use of a virus-inactivating RNA stabilization reagent to preserve SARS-CoV-2 nucleic acids – and maintain their physical linkage (**Fig. 4**) – at room temperature for 24 hours could enable a wide of variety of research and collection protocols, including collection in diverse environments, unassisted home sampling of saliva, and safe transport of inactivated samples at ambient temperatures. We hope these approaches will inform epidemiology, diagnostics, and safe management of the environments in which humans live, move, and work.

## Data Availability

Data for this study is included in figures and tables in the manuscript.

## Acknowledgements

This work was supported by the Harvard Medical School Department of Genetics. We thank Angela Reid (Harvard Environmental Health and Safety) for helpful advice, and Christopher Patil for comments on the manuscript.

